# Clinical and health system factors associated with antiretroviral therapy adherence among people living with HIV and AIDS: cross-sectional survey insights from three ART facilities in Tamale, Ghana

**DOI:** 10.1101/2025.05.28.25328495

**Authors:** Faisal Gunu Abdul-Samed, Umar Haruna, Abdulai Alimatu, Gifty Apiung Aninanya

## Abstract

HIV and AIDS have attained a manageably chronic status as more infected people experience increases in their life quality and expectancy. Health system factors affect adherence to ART among persons living with HIV and AIDS(PLHIV). Nonetheless, literature on health-system factors and ART adherence among PLHIV is sparse. This study therefore, aimed to elucidate clinical and health system factors associated with antiretroviral therapy adherence among people living with HIV and AIDS in Tamale Metropolis. We conducted a facility-based cross-sectional survey of 418 PLHIV in the Tamale Metropolis, selected by consecutive sampling from three major antiretroviral therapy (ART) centres. Each factor associated with ART adherence, considered statistically significant at a p-value of <0.05 with a 95% confidence interval, was analysed using both binary and multivariate logistic regression. The ART adherence rate was 93%. Clinical and health system factors significantly associated with higher adherence included the absence of post-pill fatigue (AOR = 0.09; 95% CI: 0.02–0.37), absence of complaints regarding pill size (AOR = 3.71; 95% CI: 1.23–11.18), lower cost of accessing therapy (AOR = 0.27; 95% CI: 0.10–0.73), uninterrupted ART supply (AOR = 7.76; 95% CI: 1.02–59.30), and strong social support systems (AOR = 6.62; 95% CI: 1.18–37.21). In conclusion, a suboptimal adherence rate of 93% was obtained. Clinical factors promoting adherence included the absence of fatigue and concerns related to pill size, while health system related factors promoting adherence included reduced cost of access, consistent ART supply, and good social support. The Ghana AIDS Commission and its implementing partners are urged to strengthen community-based social support networks, expand ART distribution points, and develop targeted educational initiatives to improve therapy adherence and contribute to achieving epidemic control.

**CONTRIBUTIONS TO THE LITERATURE:**

- Research on the clinical and health system issues associated with ART adherence among people living with HIV and AIDS in Ghana’s Northern Region is scarce, and this study helps address this gap in the literature.
- This study provides insights into individual adherence factors among PLHIV in the Tamale Metropolis, which can inform the Ministry of Health and Ghana Health Service guidance on ART adherence.
- Knowledge generated from this research can inform subnational and national policies to support PLHIV.

## 1. INTRODUCTION

Almost three decades after the 1996 introduction of the Highly Active Antiretroviral Therapy (HAART), HIV and AIDS have become manageable chronic conditions as more infected people experience increases in the quality and expectancy of their lives [1]. However, HIV and AIDS remain paramount global public health challenges, with a worldwide imperative to eradicate AIDS and accelerate the HIV response by 2030 [2]. In 2021, the incidence of newly diagnosed HIV infections was 1.5 million, with 650,000 deaths attributed to AIDS, and an approximate total of 38.4 million people living with HIV (PLHIV; 54% women and girls). Of these, 28.7 million (75% of PLHIV) could access ART. Despite efforts to curb novel infections and fatalities, Sub-Saharan Africa maintains the highest HIV prevalence globally, with approximately one in every twenty-five adults infected, constituting over two-thirds of all PLHIV worldwide. Moreover, the region is responsible for 60% of new HIV infections and half of all AIDS-related deaths globally [3,4].

Adherence is characterized as the degree to which a patient follows the instructions of a healthcare professional regarding prescribed medications [5]. Individuals embarking on ART should possess the capability and willingness to adhere to treatment, grasp both the advantages and limitations of the regimen, and recognize the significance of adherence, but patients and healthcare professionals experience additional adherence issues resulting from the switch to combination treatments for PLHIV [6]. Understanding the factors affecting PLHIV’s effective and regular adherence to ART is crucial to addressing suboptimal adherence. Clinical factors noted in the literature include the use of traditional and herbal medicine, doubts concerning the benefits of ART, side effects, and the burden of pill consumption [7–10]. Health systems factors include discontentment with healthcare facilities and staff in particular, extended wait times, stockouts/shortages, distance, unforeseen facility charges, out-of-pocket expenses for medications and transportation to services, expenses of forgoing daily income-generating activities [5,7,9,11,12]. Studies conducted in Ghana that incorporated health system determinants of ART adherence have reported adherence rates of 51.2%, 44.6%, and 73%, respectively. Key factors associated with suboptimal adherence included complex dosing regimens (i.e., more than one dose per day), the presence of treatment-related side effects, lack of regular reminders from healthcare providers, and long travel distances to ART centers, specifically distances of 51 kilometers or more [5,8,13].

Ghana currently implements a triple therapy regimen for PLHIV, including either one NtRTI, one NRTI, and one INSTI (primarily dolutegravir) as preferred first-line treatment for adults, or one NtRTI, one NRTI, and one NNRTI (primarily efavirenz) as an alternative first-line option [14]. Ghana has an estimated 342,307 PLHIV in a total population of around 31 million as of 2019. Its HIV prevalence of 1.7% suggests a moderate HIV burden compared to several African nations. However, only 46% of PLHIV receive ART [7]. AIDS-related mortality in Ghana accounts for 13,616 deaths annually [14]. In Ghana’s Northern region, HIV prevalence is 0.6%, affecting around 6,941 PLHIV. Tamale Metropolis contributes significantly, constituting 30% of the region’s prevalence and achieving 88% ART coverage [12]. While most studies conducted in Northern Ghana have explored the determinants of adherence to antiretroviral therapy (ART) [15,16], there remains a paucity of evidence specifically examining health system-related factors influencing ART adherence within healthcare facility settings

This study thus aimed to examine the clinical and health system challenges that PLHIV encounter in maintaining consistent adherence to their treatment regimen in the Tamale Metropolis. This study follows an earlier one that reported on individual and socio-cultural factors influencing adherence to ART. Objectives were to: (i) identify ART adherence rate and reported clinical and health system challenges; (ii) analyse associations between clinical and health system factors and ART adherence; and (iii) discuss implications for healthcare providers, policymakers, community organizations, or researchers working with PLHIV in Tamale Metropolis or beyond.

## MATERIALS AND METHODS

### 2.1 Study design and setting

We conducted a facility-based cross-sectional survey in three ART centres within Tamale Metropolis, namely Tamale Teaching Hospital (TTH), Tamale West Hospital (TWH), and Tamale Central Hospital (TCH). These facilities function as crucial referral points for the five northern regions of Ghana and neighbouring countries such as Togo, Ivory Coast, and Burkina Faso [17].

### 2.2 Study population

Eligibility criteria included PLHIV, aged 18 years or more, receiving ART at TTH, TWH, or TCH for at least six months, consenting to participate, and being in good physical/mental health, as determined through objective observation/interaction and a basic physical assessment. PLHIV experiencing critical clinical conditions during data collection or with speech or hearing impairments were excluded.

### 2.3 Sampling Size Determination

To estimate the desired effect and ensure the study was adequately powered, we calculated the sample size using the Cochran formula [18]:

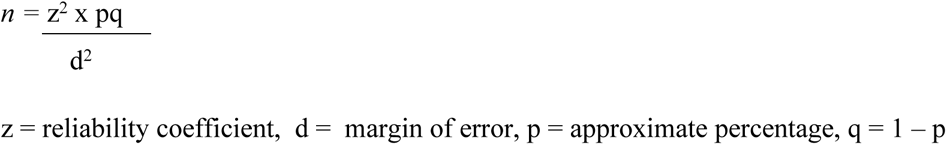

This calculation accounted for a reliability coefficient of 1.96, a 95% confidence interval (95% CI), and 80% statistical power, assuming an approximate percentage of 44.6% for PLHIV [8] and a margin of error of 5%. The estimated sample size was calculated using the following approach:

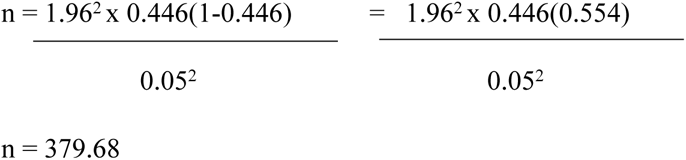

The resulting sample size was 380 participants. To address potential non-response, a 10% adjustment was applied (10% of 380 = 38), increasing the required sample size to 418 participants.

### 2.4 Sampling Techniques and procedures

Three ART facilities (TTH, TCH and TWH) were purposively selected because they are the major referral hospitals within the Tamale Metropolis. Participants were then allocated to each facility using proportional allocation based on the formula: n_i_ = (N_i_/N) x n where n_i_ is the number of participants sampled from facility I, N_i_ is the total number of eligible individuals at facility I, N is the total eligible population across all three sites, and n is the total sample size. this approach yielded 200 participants from TTH, 118 participants from TCH and 100 participants from TWH. Within each facility, convenience sampling was then employed to recruit participants who met the eligibility criteria. If a chosen individual declined to participate, another eligible participant was recruited in their place to ensure the target sample size was maintained.

### 2.5 Data collection

We adapted a questionnaire from various studies [8,17, 19–23] and pre-tested with a convenience sample of 3 PLHIV at the Tamale Central Hospital to ensure clarity and relevance. Topics included ART adherence (i.e. missed none, <3 doses, 3-12 doses, over 12 doses), socio-demographics, clinical and health system factors with scaled responses (i.e. “Strongly Agree,” “Agree,” “Disagree,” “Strongly Disagree”).

FGAS conducted structured interviews on 418 participants lasting at most 45 minutes along with 7 trained enumerators from 27/11/2023 to 20/02/2024. All were able to translate into Dagbani for participants who could not speak English. Privacy was ensured through meeting in a private room in each facility. After obtaining their consent, confidentiality was ensured by assigning all participants an identification code and not including personally identifiable data in the questionnaire or outputs. Data reliability and completeness were checked and corrected daily with Microsoft Excel used for entry of data entry and coding.

### 2.5 Analysis

Data were transferred to SPSS version 21 for analysis. The primary outcome variable was self-reported ART adherence, with a binary categorisation of ‘good adherence’ if participants reported taking at least 95% (i.e. missed none or <3 doses) of their prescribed medication in the past 30 days or ‘poor adherence’ if they reported taking less than 95% (i.e. missing 3+ doses) of their prescribed medication in the past 30 days.

Independent variables were clinical or health system-related. These were recoded to facilitate binary logistic regression, with “Strongly Agree” and “Agree” taken as Agree, while “Strongly Disagree” and “Disagree” were taken as Disagree. Descriptive statistics, including frequencies and percentages, were calculated for all variables to present a comprehensive summary of the dataset. To identify factors significantly associated with ART adherence, bivariate logistic regression was initially conducted. Variables achieving a significance level of 0.05 in the bivariate analysis were subsequently included in the multivariable logistic regression model to control for potential confounding factors. The variables included in the multivariable model were: fatigue, too many pills, pill odour, auditory hallucination, pill size, cost of accessing ART, social support, interrupted ART supply and cost of treating comorbidities. These variables were selected based on their significance in the bivariate analysis.

To ensure the validity of the responses related to variables influencing ART adherence, a pre-assessment of the internal consistency of the survey instrument was conducted using Cronbach’s Alpha analysis. This method assessed the reliability of participants’ responses across the 31 survey items. A Cronbach’s Alpha coefficient of 0.8 was obtained, demonstrating good internal consistency. This step, conducted before analyzing the data, confirmed the reliability of the instrument in capturing meaningful responses about ART adherence.

## 2. RESULTS

### 3.1 Participant characteristics

The demographic profile of the 418 PLHIV surveyed shows a predominance of females (74.4%), most were between the ages of 30–49 years (58.6%), married or cohabiting (65.3%), Muslim (71.5%), and of Dagomba ethnicity (64.6%), reflecting the local socio-cultural landscape. Educational attainment was low, with over one-third (35.4%) having no formal education whilst majority (73%) lived in urban areas.

**Table 1:** Demographic Characteristics of PLHIV on ART.

| **Variable** | **Tamale**<br>**Central**<br>**(n=118)** | **Tamale**<br>**West**<br>**(n=100)** | **Tamale**<br>**Teaching**<br>**(n=200)** | **Total**<br>**(N=418)** |
| --- | --- | --- | --- | --- |
| **Age Groups** |  |  |  |  |
| 18-29 | 23 (19.5%) | 25 (25.0%) | 51 (25.5%) | 99 (26.3%) |
| 30-49 | 76 (64.4%) | 59 (59.0%) | 104 (52.0%) | 239 (58.6%) |
| ≥50 | 19 (16.1%) | 16 (16.0%) | 45 (22.5%) | 80 (15.1%) |
| **Gender** |  |  |  |  |
| Male | 27 (22.9%) | 20 (20.0%) | 60 (30.0%) | 107 (25.6%) |
| Female | 91 (77.1%) | 80 (80.0%) | 140 (70.0%) | 311 (74.4%) |
| **Marital Status** |  |  |  |  |
| Married/Cohabiting | 73 (61.9%) | 61 (61.0%) | 139 (69.5%) | 273 (65.3%) |
| Single | 24 (20.3%) | 31 (31.0%) | 38 (19.0%) | 93 (22.3%) |
| Widowed/Divorced | 21 (17.8%) | 8 (8.0%) | 23 (11.5%) | 52 (12.4%) |
| **Religion** |  |  |  |  |
| - Islam | 85 (72.0%) | 79 (79.0%) | 135 (67.5%) | 299 (71.5%) |
| - Christianity | 33 (28.0%) | 21 (21.0%) | 65 (32.5%) | 119 (28.5%) |
| **Education Level** |  |  |  |  |
| - No Formal Education | 40 (33.9%) | 41 (41.0%) | 67 (33.5%) | 148 (35.4%) |
| - Formal Education | 78 (66.1%) | 59 (59.0%) | 133 (66.5%) | 270 (64.6%) |
| **Settlement Type** |  |  |  |  |
| - Urban | 80 (67.8%) | 86 (86.0%) | 139 (69.5%) | 305 (73.0%) |
| - Rural/Suburban | 38 (32.2%) | 14 (14.0%) | 61 (30.5%) | 120 (29.0%) |
| **Tribe** |  |  |  |  |
| - Dagomba | 80 (67.8%) | 70 (70.0%) | 120 (60.0%) | 270 (64.6%) |
| - Mamprusi | 10 (8.5%) | 3 (3.0%) | 21 (10.5%) | 34 (8.1%) |
| - Others* | 28 (23.7%) | 27 (27.0%) | 59 (29.5%) | 114 (27.3%) |

*Note.* Others\*: Mossi, Gonja, Frafra, Builsa, Waala, Kasina, Ga, Dagaati, Fante, Ashanti, Ewe, Hausa, Bimoba, Zabarma. Data were collected from three hospitals in the Tamale Metropolis (Ghana): Tamale Teaching Hospital, Tamale Central Hospital, and Tamale West Hospital. The study period was from November 2023 to February 2024.

### 3.2 ART adherence

Among the 418 PLHIV surveyed, 93.1% demonstrated good adherence (defined as ≥95% of prescribed doses taken in the past 30 days). The highest adherence was recorded at Tamale Teaching Hospital (95.5%), compared to Tamale West Hospital (89%) and Tamale Central Hospital (92.4%).

**Table 2:** ART Adherence Rates.

| Facility | Good Adherence | Poor Adherence | Total |
| --- | --- | --- | --- |
| Tamale Central Hospital | 109 (92.4%) | 9 (7.6%) | 118 |
| Tamale West Hospital | 89 (89.0%) | 11 (11.0%) | 100 |
| Tamale Teaching Hospital | 191 (95.5%) | 9 (4.5%) | 200 |
| **Overall** | 389 (93.1%) | 29 (6.9%) | 418 |

*Note.* Data were collected from three hospitals in the Tamale Metropolis (Ghana): Tamale Teaching Hospital, Tamale Central Hospital, and Tamale West Hospital.

### 3.3 Potential clinical factors affecting ART adherence

Table 3 shows that while most participants did not report any issues with their pills or challenging side effects, 69% reported they would prefer an injectable ART. Most (58%) also reported adhering to their ART regimen despite experiencing side effects.

**Table 3.**
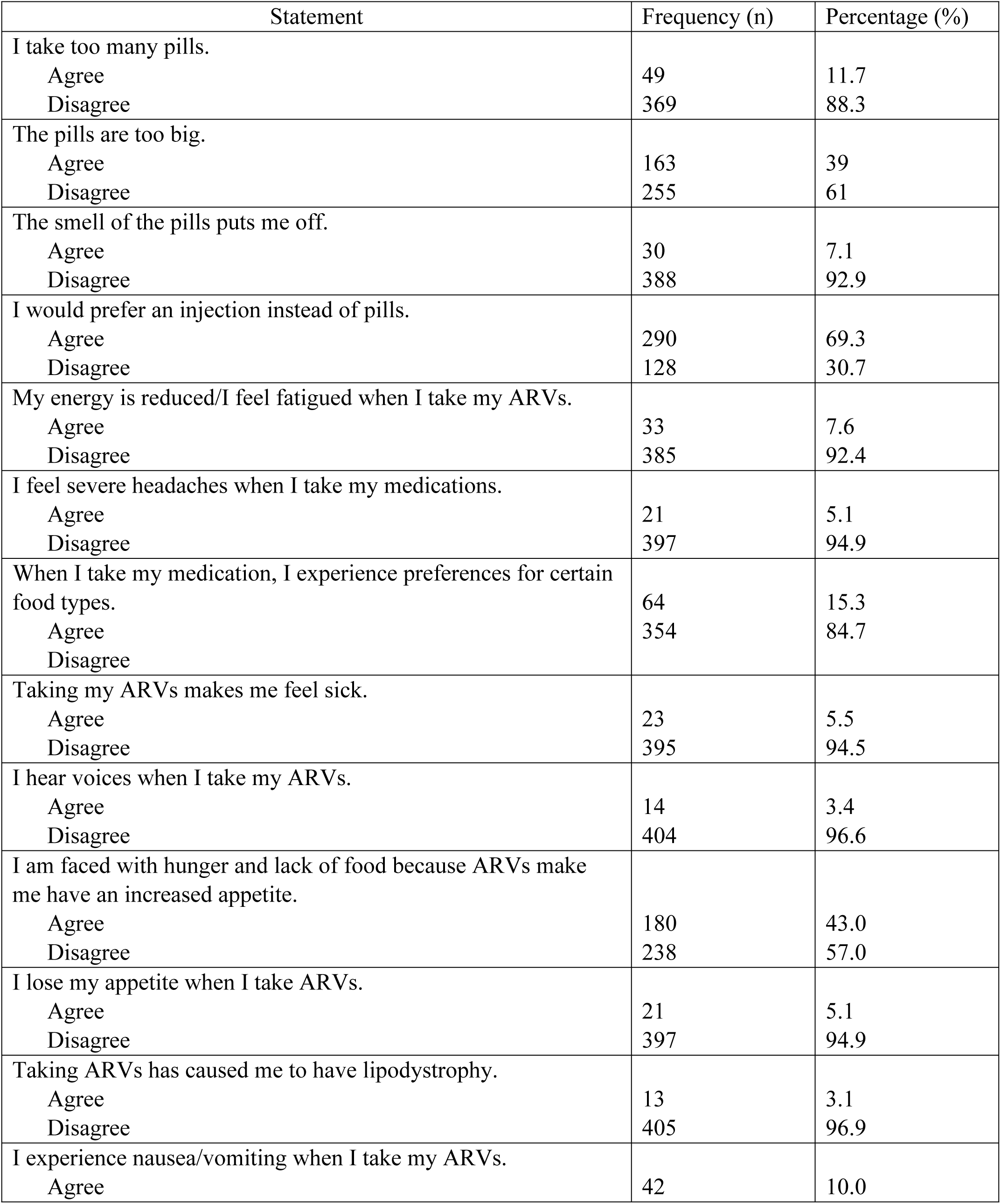
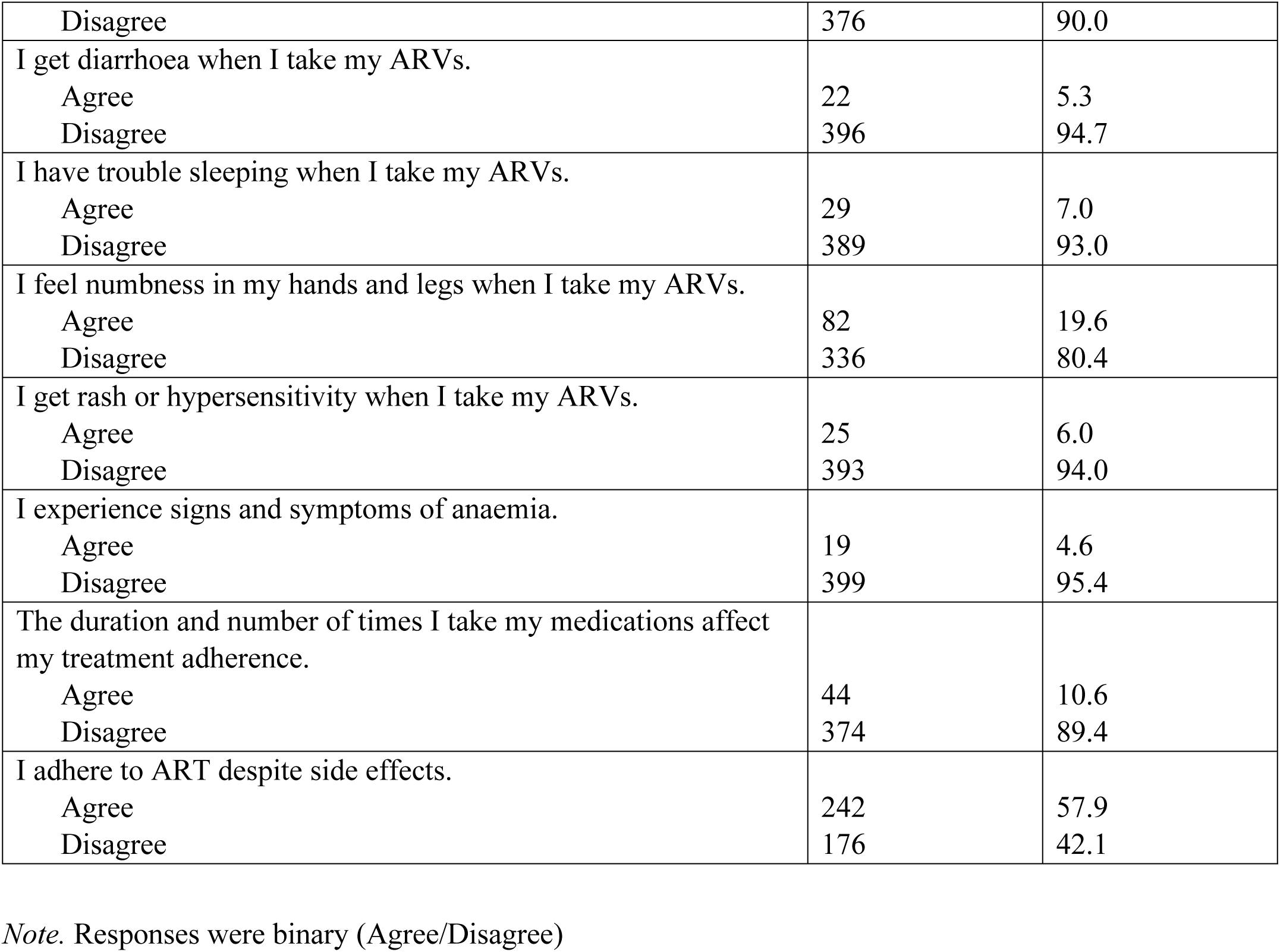
Medication-related factors affecting ART adherence.

### 3.4 Potential Health system Factors Affecting ART adherence

Table 4 shows most participants reported most health system factors positively. For example, 91% reported favourable health-worker influence, 95% reported that health workers emphasized the importance of medication adherence, and 70% received refill reminders. Over 95% reported not experiencing any harsh treatment, discrimination, or abuse at healthcare facilities. Most indicated ART services were always available and accessible (98%), were confident in facility confidentiality practices (91%), and experienced acceptable financial (84%) and geographical (65%) barriers. Notably, only 22% reported receiving social support from NGOs, welfare units, or volunteers. The highest negative response was 55% reporting that increasing patient numbers that made it more crowded to access ART at their facility impacted their adherence.

**Table 4.** Health System Factors Affecting ART Adherence.

| Statement | Frequency (n) | Percentage (%) |
| --- | --- | --- |
| I was shouted at/treated harshly by healthcare providers.<br>Agree<br>Disagree | 18<br>400 | 4.3<br>95.7 |
| Health workers took me through the importance of taking my medications.<br>Agree<br>Disagree | 395<br>23 | 94.5<br>5.5 |
| I have a cordial relationship with my healthcare providers.<br>Agree<br>Disagree | 382<br>36 | 91.4<br>8.6 |
| I experience discrimination and abuse in the healthcare unit.<br>Agree<br>Disagree | 22<br>396 | 5.3<br>94.7 |
| I face high financial costs when accessing and receiving ART (e.g. transport, cost of drugs).<br>Agree<br>Disagree | 68<br>350 | 16.3<br>83.7 |
| Interrupted ART supply from healthcare provider (e.g. due to a shortage of drugs and other commodities).<br>Agree<br>Disagree | 34<br>384 | 8.2<br>91.8 |
| It takes lots of time for a refill of my medications (long waiting time).<br>Agree<br>Disagree | 26<br>392 | 6.2<br>93.8 |
| I lose my income on ART therapy including treating underlying diseases/infections.<br>Agree<br>Disagree | 12<br>406 | 2.9<br>97.1 |
| The distance from where I stay to the ART centre influences my adherence.<br>Agree<br>Disagree | 148<br>270 | 35.4<br>64.6 |
| I get reminders from health workers for a refill (positive patient-provider partnership).<br>Agree<br>Disagree | 292<br>126 | 69.9<br>30.1 |
| The ART services are always available and easily accessible.<br>Agree<br>Disagree | 409<br>9 | 97.8<br>2.2 |
| I receive social support (e.g. from the welfare unit, government, NGO, volunteers).<br>Agree<br>Disagree | 91<br>327 | 21.8<br>78.2 |
| There is a lack of confidentiality/respect for privacy at the facility.<br>Agree<br>Disagree | 40<br>378 | 9.5<br>90.5 |
| The number of people accessing ARVs is always increasing at this hospital.<br>Agree<br>Disagree | 230<br>188 | 55.0<br>45.0 |
| Health workers are very slow and not organized because when you come no matter how early, you will still leave there late.<br>Agree<br>Disagree | 28<br>390 | 6.7<br>93.3 |

*Note.* Responses were binary (Agree/Disagree)

### 3.5 Clinical and Health System-related Determinants of Adherence to ART

After adjusting for potential confounding variables using multivariable logistic regression analysis, several factors remained significantly associated with adherence to antiretroviral therapy (ART). These included both clinical and health system-related determinants. Clinically, the absence of post-pill fatigue and the lack of concern regarding pill size were positively associated with adherence. From a health system perspective, reduced financial burden, uninterrupted ART supply, and access to strong social support whether from NGOs, government programs, or community networks also significantly enhanced adherence.

Specifically, participants who did not experience post-pill fatigue had 91% lower odds of non-adherence compared to those who did (AOR = 0.09; 95% CI: 0.02–0.37). Similarly, individuals who had no issues with pill size were 3.71 times more likely to adhere to treatment than those who found pill size problematic (AOR = 3.71; 95% CI: 1.23–11.18). Those reporting less financial strain in accessing therapy were 73% more likely to adhere compared to participants experiencing higher financial burden (AOR = 0.27; 95% CI: 0.10–0.73). Moreover, uninterrupted ART supply significantly improved adherence, with such individuals being 7.76 times more likely to adhere (AOR = 7.76; 95% CI: 1.02– 59.30). Finally, strong social support was a robust predictor of adherence, as participants with support networks were over six times more likely to remain adherent than those without (AOR = 6.62; 95% CI: 1.18–37.21). The combination of these five variables explained 16.4% of the variance in ART adherence (Nagelkerke R² = 0.164). The model demonstrated good fit to the data, as indicated by a non-significant Pearson Chi-Square test (χ² = 29.51, df = 22, p = 0.131) and Deviance statistic (χ² = 24.45, df = 22, p = 0.324), suggesting that the predicted values from the model do not significantly differ from the observed values, supporting the model’s adequacy in explaining ART adherence behavior.

**Table 5.**
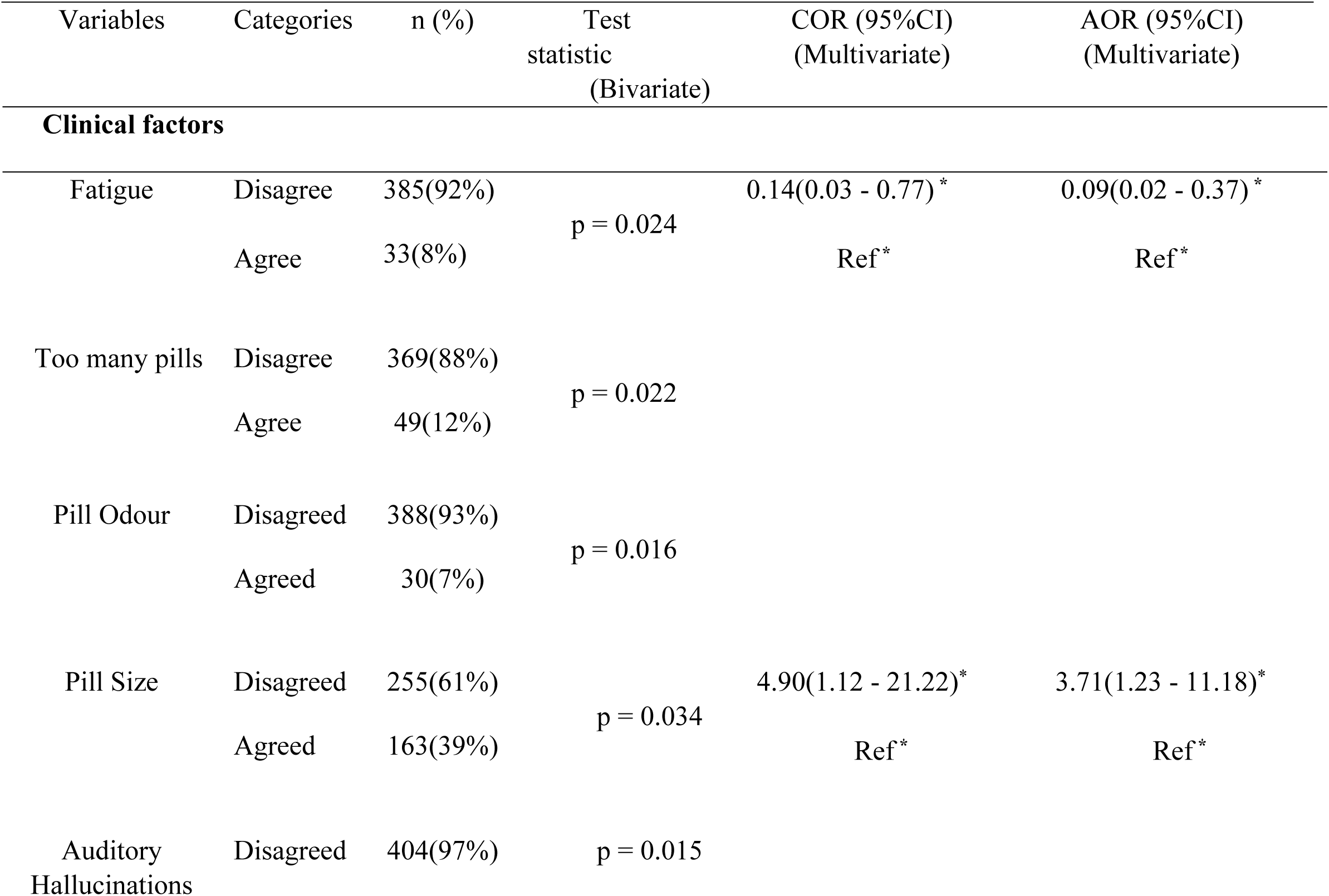
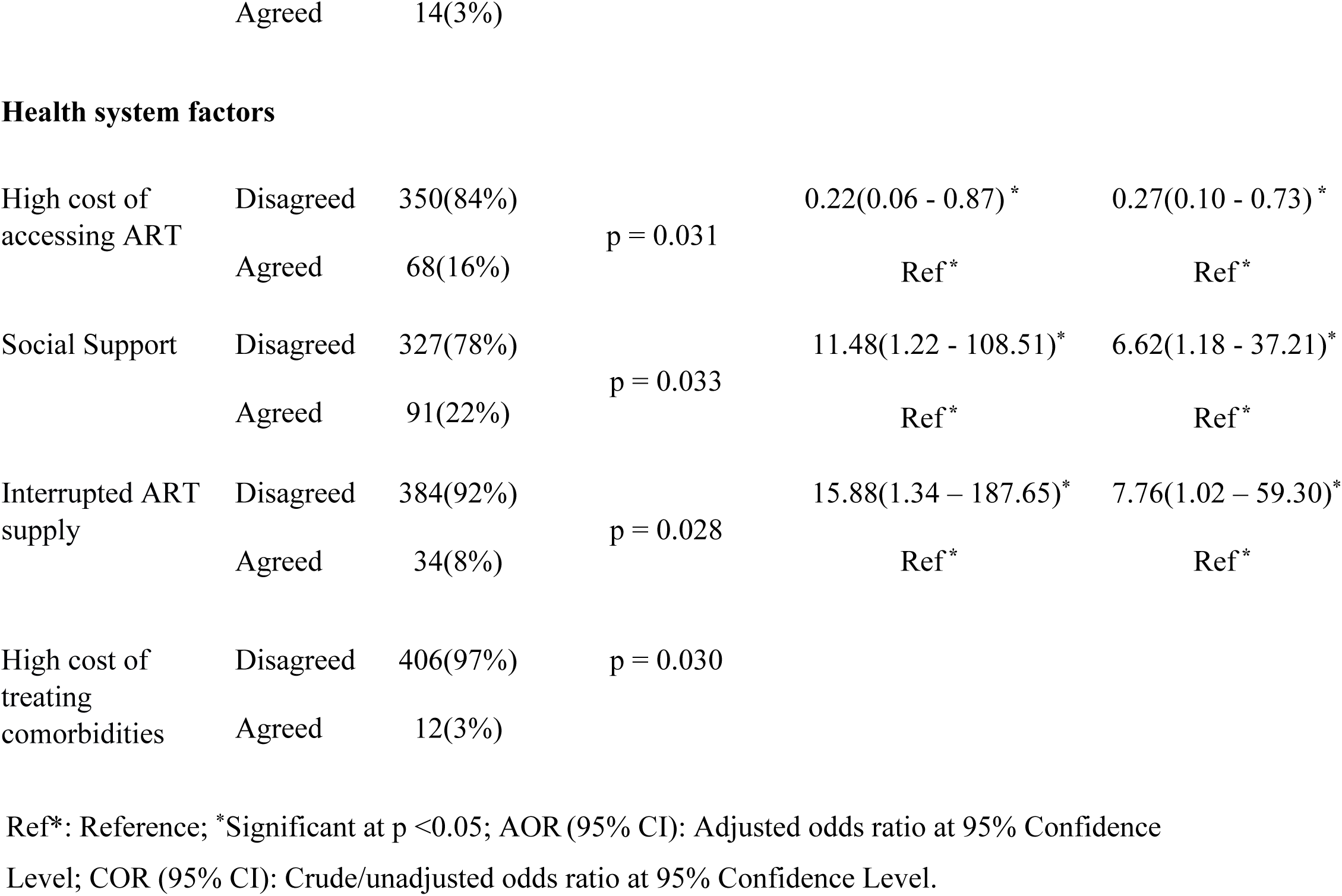
Determinants of Adherence to ART among PLHIV.

## 4. DISCUSSION

This study is one of the first to explore the relationship between clinical and health system factors and ART adherence in Northern Ghana, offering vital contextual insights. The reported adherence rate of 93%, though encouraging, falls marginally below the 95% benchmark set by UNAIDS for achieving sustained viral suppression and halting the global spread of HIV/AIDS by 2025 [15,24]. Globally, ART adherence remains inconsistent, with reported rates ranging from 49% to 100% [25]. The adherence level observed in this study closely mirrors findings from Nigeria (92.6%) [26], suggesting some consistency in ART uptake across West African contexts. However, it contrasts significantly with lower adherence rates reported in other parts of Ghana such as the Sunyani Municipality (75%), Ga West Municipality (44.6%), and Cape Coast Metropolis (79.5%) [8,16]. These discrepancies may reflect regional variations in health system capacity, socio-economic factors, and community engagement in HIV care. The suboptimal adherence rate observed in this study suggests that when adequately resourced and contextually adapted, ART programs in Northern Ghana can meet or approach global benchmarks. This reinforces the need to replicate and scale context-specific adherence strategies across regions with suboptimal performance.

Clinically, the study identified post-pill fatigue and pill size as significant barriers to adherence. While pill size is not a physiological side effect, it emerged as a perceptual barrier impacting patient motivation to maintain treatment regimens. These findings are consistent with earlier studies which highlighted how both physiological side effects and formulation-related inconveniences undermine adherence [21, 24, 27–31]. O’Connell et al. (2022) noted that the emergence of side effects during ART initiation often leads to therapy interruptions, while Loveday et al. (2022) emphasized that any adverse effect impairing a patient’s functional capacity poses a long-term threat to adherence. These findings highlight the importance of ongoing pharmacovigilance, patient-centered drug formulation (e.g., smaller or more palatable pills), and early side effect management. Health facilities should incorporate routine screening and counseling for side effects as part of ART adherence interventions.

Financial access emerged as another critical determinant. Participants with reduced financial burden were 73% more likely to adhere to treatment. This aligns with findings by Loveday et al. (2022), who emphasized the economic pressures such as employment retention and transport costs as barriers to ART adherence. These results reinforce the importance of policies that reduce out-of-pocket expenses, such as subsidized ART-related services or transport support. Financial protection mechanisms must be integrated into national HIV programs to improve long-term treatment retention.

Social support was shown to significantly enhance adherence, increasing the likelihood of consistent treatment by more than sixfold. This corroborates findings from prior studies which reported that peer groups, supportive families, and stigma-free communities are strongly associated with improved adherence outcomes [23, 26, 28, 32–35]. Conversely, social isolation or stigma can fracture critical support systems. HIV care programs must embed psychosocial interventions including peer support groups and community sensitization campaigns into ART delivery models. Tailored support systems can buffer stigma and reinforce treatment commitment.

Health system functionality, particularly uninterrupted ART supply, also proved critical. Individuals with consistent access were 7.76 times more likely to adhere, echoing studies that have reported drug stock-outs, clinic appointment delays, and occupational conflicts as major adherence barriers [26,32]. Strengthening ART supply chains and enhancing health facility responsiveness are non-negotiable. Integration of digital inventory tracking systems, improved staff capacity, and workplace flexibility policies for ART clients should be prioritized.

Collectively, these findings carry critical implications for clinical practice, health policy, and HIV program design. They underscore the need for comprehensive, multi-level interventions that address not only the clinical dimensions of HIV care but also the socio-economic and systemic determinants of adherence. Key strategies include ensuring the consistent availability of ART, minimizing patient costs, proactively managing side effects, and fostering robust community support structures. The involvement of civil society organizations, community health workers, and policy makers will be essential in translating these findings into sustainable programmatic improvements.

4.1 Limitations

This study offers important insights into ART adherence in Northern Ghana. The high response rate and consistent data collection procedures also enhance the reliability of the findings. Nonetheless, several limitations should be acknowledged. First, the study excluded individuals in their first six months of ART initiation, those who were critically ill, co-infected with tuberculosis, or with speech and hearing impairments. As a result, some unique adherence challenges experienced by these groups may not have been captured. Future studies should broaden inclusion criteria to reflect a more diverse patient population and ensure greater inclusivity.

Second, the cross-sectional nature of the study limits the ability to draw causal inferences between the identified factors and ART adherence. To better understand temporal dynamics and causal pathways, longitudinal and interventional studies are recommended. Third, although the study employed a consecutive sampling method, a non-probabilistic approach that limits generalizability, we mitigated potential selection bias by recruiting participants systematically as they presented at ART clinics. Additionally, data collection across multiple health facilities with diverse demographics helped to improve representativeness and reduce the likelihood of sampling skew.

Despite these limitations, the study offers valuable baseline data and contextual evidence that can inform policy and programming efforts to improve ART adherence in similar low-resource settings.

4.1 Conclusion

This study identified a suboptimal 93% ART adherence rate among PLHIV, clinical factors promoting adherence included the absence of fatigue and concerns related to pill size, while health system related factors promoting adherence included reduced cost of access, consistent ART supply, and good social support. The Ghana AIDS Commission and its implementing partners are urged to strengthen community-based social support networks, expand ART distribution points, and develop targeted educational initiatives to improve therapy adherence and contribute to achieving epidemic control.

## DECLARATIONS

### Author Contributions

Conceptualization and Methodology: FGAS, and GAA; Validation: GAA; Formal analysis, resources and data collection: FGAS; Writing of the manuscript: FGAS, GAA, A.A and UH.

### Ethics approval

Ethics committees at the Tamale Teaching Hospital (references TTHERC/20/11/23/01) and Ghana Health Service (GHS-ERC:053/09/23) provided ethics approval respectively.

### Informed Consent Statement

Every participant in the study provided written, informed consent.

### Data Availability

Upon a reasonable request, the corresponding author can supply the data sets used in the current work.

## Abbreviations

ART: Antiretroviral Therapy
PLHIV: People Living with HIV
HIV: Human Immunodeficiency Virus
AIDS: Acquired Immunodeficiency Syndrome
HAART: Highly Active Anti-Retroviral Therapy
TTH: Tamale Teaching Hospital
TWH: Tamale West Hospital
TCH: Tamale Central Hospital

## Acknowledgements

The authors thank the study participants for their time and hospital workers and management of the ART/STI clinic for their support.

